# Weight-Loss Efforts in US Adults with Hypertension and Overweight or Obesity

**DOI:** 10.64898/2026.03.23.26349132

**Authors:** Guang Xiong, Ruimin Tian, Michelle Shukhman, John Andraos, Yijun Cai, Junjie Lu, Heqing Tao, Ligang Liu

## Abstract

**Background:** Obesity has become increasingly common among US adults with hypertension. However, national data are limited on weight-loss efforts among adults with hypertension and overweight/obesity, and whether these efforts have translated into clinically meaningful weight loss at the population level.

**Methods:** We analyzed repeated cross-sectional data from the National Health and Nutrition Examination Survey, 1999-2023. Adults aged ≥20 years with hypertension and body mass index ≥25 kg/m² were included. Weight-loss attempt was defined as self-report of trying to lose weight during the prior 12 months. Among those attempting weight loss, successful weight loss was defined as ≥5% or ≥10% reduction in body weight over the prior year. Survey-weighted logistic regression was used to assess temporal trends and associations between strategies and successful weight loss.

**Results:** Overall, 57.6% reported a weight-loss attempt, increasing from 55.9% in 1999-2000 to 60.4% in 2021-2023 (P for trend=0.002). The most reported strategies were eating less food (65.3%) and exercise (52.4%). Among those attempting weight loss, 33.4% achieved ≥5% weight loss and 14.7% achieved ≥10% weight loss; neither improved over time (P for trend=0.976 and 0.174, respectively). Weight-loss surgery was strongly associated with success but was rarely reported (0.35%). Eating less fat and changing eating habits were also positively associated with successful weight loss, whereas skipped meals and use of diet foods or products were inversely associated.

**Conclusions:** Weight-loss attempts increased, but clinically meaningful weight-loss success did not improve, highlighting a persistent gap between effort and outcome in hypertension care.

## 1. INTRODUCTION

The concurrent epidemics of obesity and hypertension present a major public health challenge. Among US adults with hypertension, the prevalence of obesity has risen dramatically, from 39.6% in 2001 to 55.4% in 2023 [1]. Excess adiposity is not only a comorbidity but also a major pathogenic driver of hypertension through sympathetic nervous system overactivation, activation of the renin-angiotensin-aldosterone system, renal sodium retention, and vascular dysfunction [2, 3]. As a result, obesity in adults with hypertension is associated with more difficult blood pressure control, greater cardiometabolic risk, and increased target organ damage [4].

Weight loss is therefore a central component of hypertension management. Clinically meaningful weight reduction improves blood pressure, glycemic control, and lipid profiles and lowers overall cardiovascular risk [5, 6]. In adults with excess weight, each 1-kg reduction in body weight is associated with an average 1-mm Hg decrease in systolic blood pressure [7]. Accordingly, major guidelines recommend weight management as a foundational strategy for blood pressure reduction and cardiovascular disease prevention [8–10]. The 2025 American Heart Association/American College of Cardiology (AHA/ACC) guidelines identifies weight management as a core strategy for improving cardiometabolic health [8], and international guidelines similarly emphasize weight control alongside sodium reduction and physical activity [9, 10].

To address obesity more effectively, professional societies have recommended a stepped-care approach to weight management [11]. For individuals who do not achieve or maintain adequate weight loss with lifestyle intervention alone, escalation to evidence-based anti-obesity pharmacotherapy or metabolic procedures is recommended because these therapies can produce larger and more sustained reductions in adiposity [12]. The emergence of highly effective anti-obesity medications has further expanded the potential to achieve weight-loss targets that were previously difficult for many patients to reach [13].

Despite these recommendations and therapeutic advances, little is known about how weight-loss efforts are being implemented among adults with hypertension at the population level. National surveillance studies have described weight-loss attempts and strategies in the general US population [14, 15], but adults with hypertension have not been specifically examined. This distinction is clinically important because weight reduction in this population is directly relevant to blood pressure control and prevention of downstream complications. In addition, changes over the past two decades in dietary guidance, public health messaging, health care delivery, and the availability of newer pharmacotherapies may have influenced weight-loss behaviors and outcomes in this group [16–18]. Whether these changes have translated into greater success in achieving clinically meaningful weight loss remains unclear.

To address these gaps, we analyzed data from the National Health and Nutrition Examination Survey (NHANES) from 1999 to 2023. To address this gap, we analyzed data from the National Health and Nutrition Examination Survey (NHANES) from 1999 to 2023 among US adults with hypertension and overweight or obesity. We sought to quantify long-term trends in weight-loss attempts, estimate the prevalence and trends of clinically meaningful weight-loss success (≥5% and ≥10%) among those attempting to lose weight, characterize self-reported weight-loss strategies, and examine the associations between specific strategies and successful weight loss.

## 2. METHODS

### 2.1 Study overview

We conducted a cross-sectional analysis of data from the National Health and Nutrition Examination Survey (NHANES) from 1999 to 2023. NHANES is a nationally representative survey of the civilian, noninstitutionalized US population that uses a complex, multistage probability sampling design. Survey data are collected through standardized in-home interviews and physical examinations performed by trained personnel in mobile examination centers. To obtain stable national estimates, we pooled data from 12 consecutive 2-year survey cycles (1999-2000 through 2021-2023). All NHANES protocols were approved by the National Center for Health Statistics Research Ethics Review Board, and all participants provided written informed consent.

The study population included adults aged ≥20 years with hypertension and overweight or obesity. Among 119,555 NHANES participants initially identified, we excluded those aged <20 years, pregnant individuals, and those without hypertension (n=91,824). Among the remaining 27,731 adults with hypertension, we further excluded those with missing body mass index (BMI) data or without overweight or obesity (n=7,344). We then excluded participants with missing self-reported current weight or weight 1 year earlier (n=91) and those with missing covariate data or sample weights (n=2,913). The final analytic sample consisted of 17,383 adults (Figure 1).

**Figure 1.**
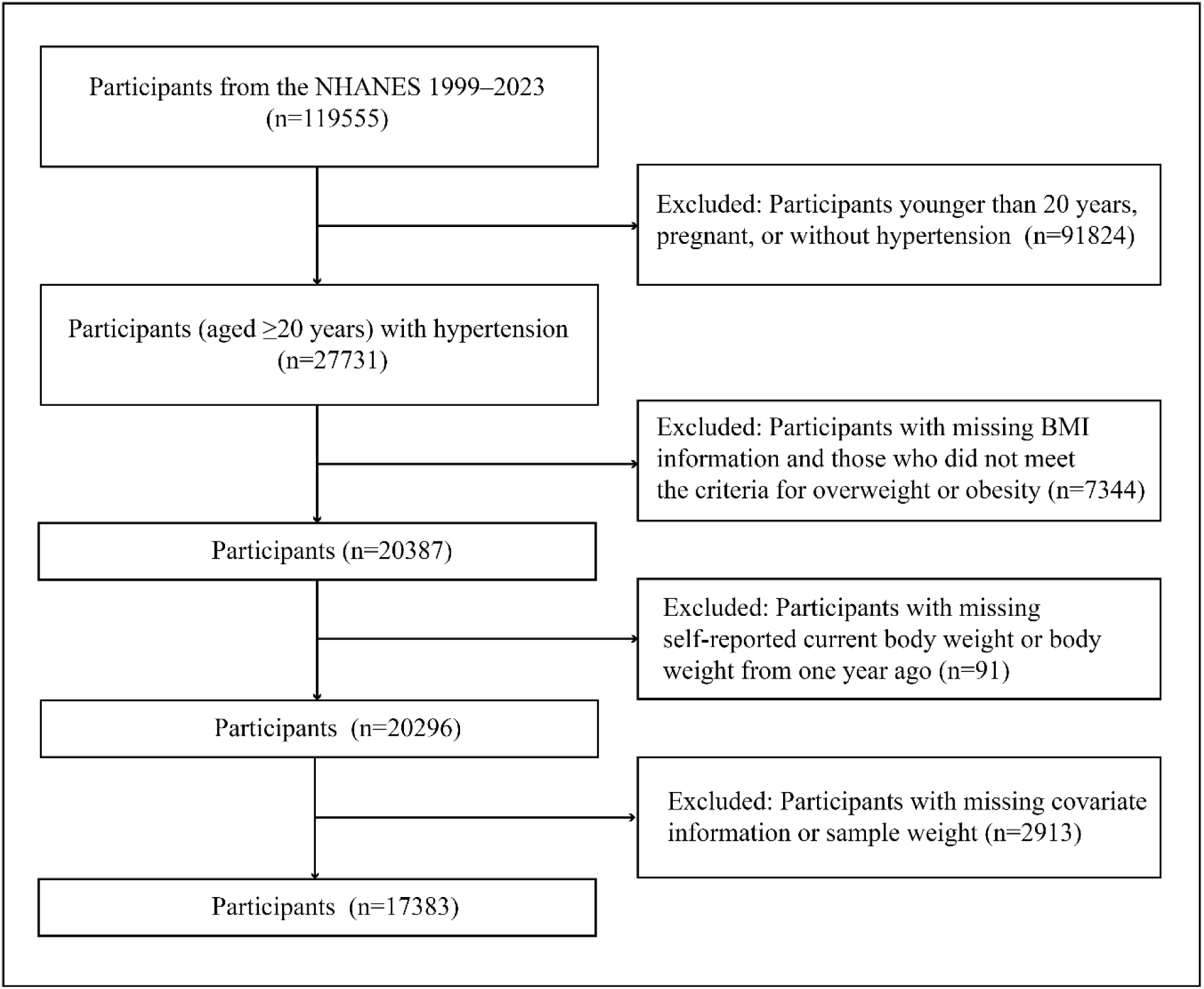
Flowchart of Participant Selection in the Study.

### 2.2 Definition of hypertension and weight status

Hypertension was defined using both examination and interview data from NHANES. Seated blood pressure was measured by trained personnel in the mobile examination center according to standardized protocols, and the mean of all available readings was used. Participants were classified as having hypertension if they had a systolic blood pressure ≥140 mmHg, a diastolic blood pressure ≥90 mmHg, self-reported physician-diagnosed hypertension, or reported use of antihypertensive medication. We used the ≥140/90 mmHg definition to maintain consistency with long-standing traditional clinical guidelines [19].

Body mass index (BMI) was calculated as measured weight in kilograms divided by measured height in meters squared (kg/m^2^). Participants were categorized as having overweight (BMI 25.0-29.9 kg/m^2^) or obesity (BMI ≥30.0 kg/m^2^) according to standard definitions [20]. BMI category was used for stratified analyses and as a covariate in regression models.

### 2.3 Weight-loss attempts, outcomes, and strategies

Weight-loss attempt was defined using the NHANES weight history questionnaire. Participants were classified as attempting weight loss if they responded affirmatively to the question, “During the past 12 months, have you tried to lose weight?”

Among participants who reported attempting to lose weight, we assessed weight change using self-reported current body weight and self-reported body weight 1 year earlier. Percent weight change was calculated as the percentage reduction from body weight 1 year earlier to current weight. Clinically meaningful weight-loss success was defined using 2 prespecified thresholds: ≥5% and ≥10% reduction in initial body weight over the preceding year. These thresholds were selected because 5% to 10% weight loss is considered clinically meaningful in current obesity and cardiovascular guidance [12, 21]. Rates of successful weight loss were calculated among participants who reported attempting to lose weight.

Weight-loss strategies were assessed among participants attempting to lose weight using a standardized NHANES checklist. Across all included survey cycles, 14 strategies were consistently assessed: ate less food, switched to foods with lower calories, ate less fat, exercised, skipped meals, used diet foods or products, used a liquid diet formula, joined a weight-loss program, took prescription diet pills, took nonprescription diet pills or supplements, used laxatives or vomited, drank a lot of water, followed a special diet, and other. Beginning in 2005-2006, 5 additional strategies were added: ate less carbohydrates, smoking, ate more fruits, vegetables, or salads, changed eating habits, and ate less sugar, candy, or sweets. “Ate less junk food or fast food” was added in 2009-2010, and “had weight-loss surgery” was added beginning in 2013-2014. Participants could report more than 1 strategy, and responses were not mutually exclusive. Because weight-loss strategy data were not collected in NHANES 2021-2023, analyses of weight-loss strategies were restricted to 1999-2020.

### 2.4 Covariates

Sociodemographic covariates were selected a priori. Age was modeled as a continuous variable. Sex was categorized as male or female. Race and ethnicity was categorized as non-Hispanic White, non-Hispanic Black, Hispanic, and other race. Educational attainment was categorized as less than high school, high school or equivalent, and college or above. Family income level was categorized as low, middle, or according to the NHANES family income-to-poverty ratio categories. BMI category (overweight versus obesity) was also included.

### 2.5 Statistical analysis

All analyses accounted for the complex, multistage probability sampling design of NHANES by incorporating strata, primary sampling units, and examination or interview weights, as appropriate. For analyses requiring measured anthropometrics and blood pressure, we used mobile examination center examination weights.

Participant characteristics were summarized as survey-weighted means with standard errors for continuous variables and as unweighted counts with survey-weighted percentages for categorical variables. We estimated the survey-weighted prevalence of weight-loss attempts overall and within prespecified subgroups defined by sex, race and ethnicity, education level, income level, and BMI category. Among participants who reported attempting to lose weight, we estimated the survey-weighted prevalence of achieving clinically meaningful weight loss at the ≥5% and ≥10% thresholds, overall and within the same subgroups.

Temporal trends in weight-loss attempts and successful weight loss (≥5% and ≥10%) across survey cycles were evaluated using survey-weighted linear or logistic regression models. All trend models were adjusted for age, sex, race and ethnicity, education level, family income level, and BMI category. For weight-loss strategies, we calculated cycle-specific weighted prevalence estimates and assessed temporal trends using similarly adjusted models, restricting each strategy analysis to survey cycles in which that item was available.

To examine associations between self-reported weight-loss strategies and successful weight loss among participants attempting to lose weight, we used survey-weighted logistic regression models with successful weight loss (≥5% or ≥10%) as the dependent variable and each strategy as a binary exposure. We prespecified 2 multivariable approaches. Model 1 included all strategy variables simultaneously. Model 2 used backward stepwise selection among strategy variables to identify a parsimonious set of strategies independently associated with success. Both models were adjusted for age, sex, race and ethnicity, education level, family income level, and BMI category. Results are reported as odds ratios (ORs) with 95% confidence intervals (CIs). Statistical significance was defined as a 2-sided P<0.05. All analyses were conducted in R (version 4.4.1) using survey packages to account for the complex sampling design.

### 2.6 Data Availability

All NHANES data used in this analysis are publicly available from the National Center for Health Statistics. Analytic documentation, including variable definitions, code, and derived measures, can be made available from the corresponding author upon reasonable request.

## 3. Results

### 3.1 Participant Characteristics

The analytic sample included 17,383 US adults aged ≥20 years with hypertension and overweight or obesity from NHANES 1999-2023 (Figure 1). The mean age was 56.2 years, 52.4% were men, and 57.3% had obesity. By race and ethnicity, 69.5% were non-Hispanic White, 14.3% were non-Hispanic Black, 5.8% were Mexican American, and 10.5% were of other races and/or ethnicities. Mean BMI was 32.1 kg/m^2^ (Table 1). Across survey cycles, several participant characteristics changed significantly. Mean age, BMI, initial weight, current weight, and 1-year weight change increased over time, and the proportion of participants with obesity increased from 55.4% in 1999-2000 to 60.4% in 2021-2023 (P<0.001). Participant characteristics stratified by BMI category are presented in Table S1.

**Table 1.**
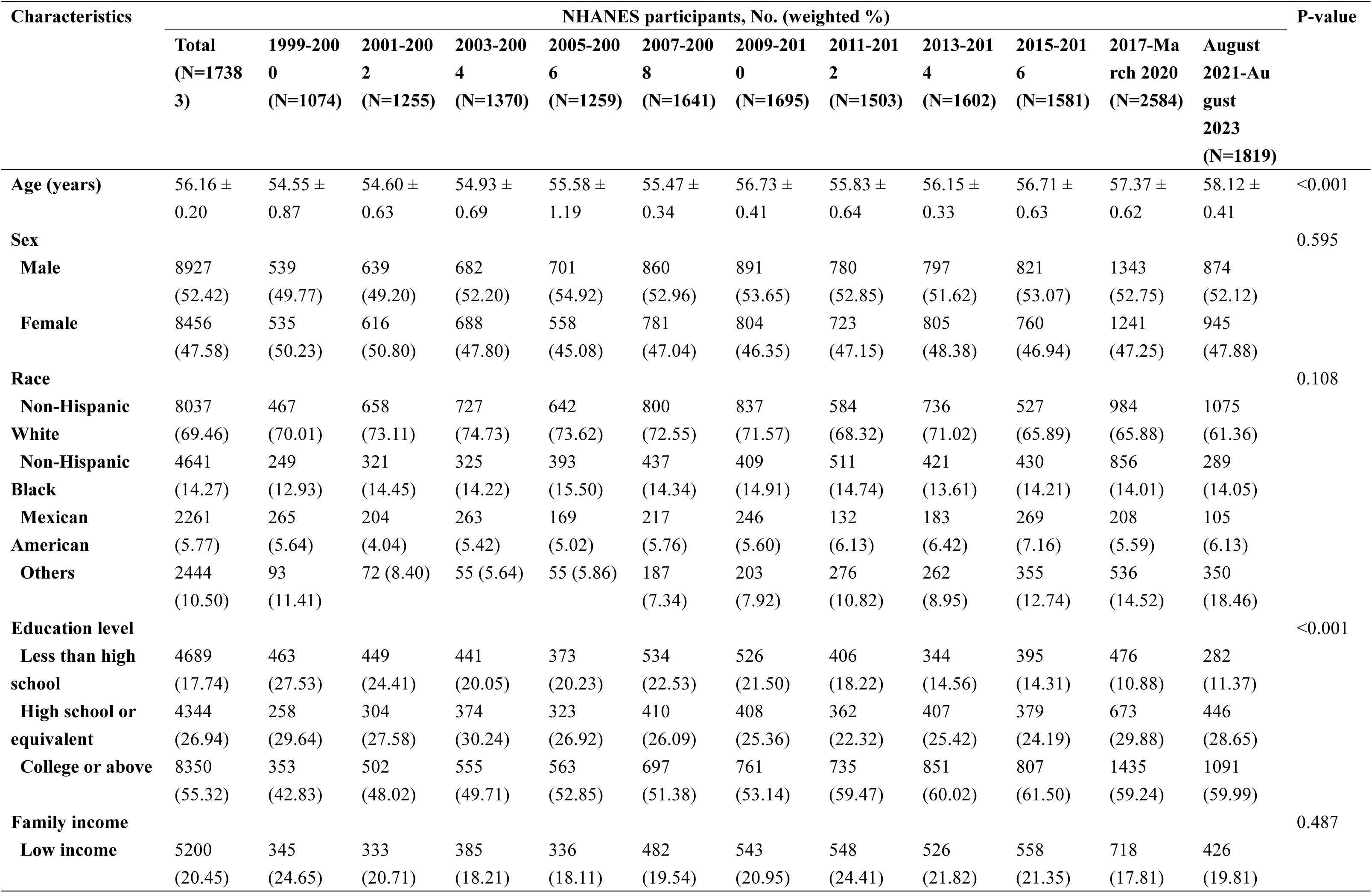
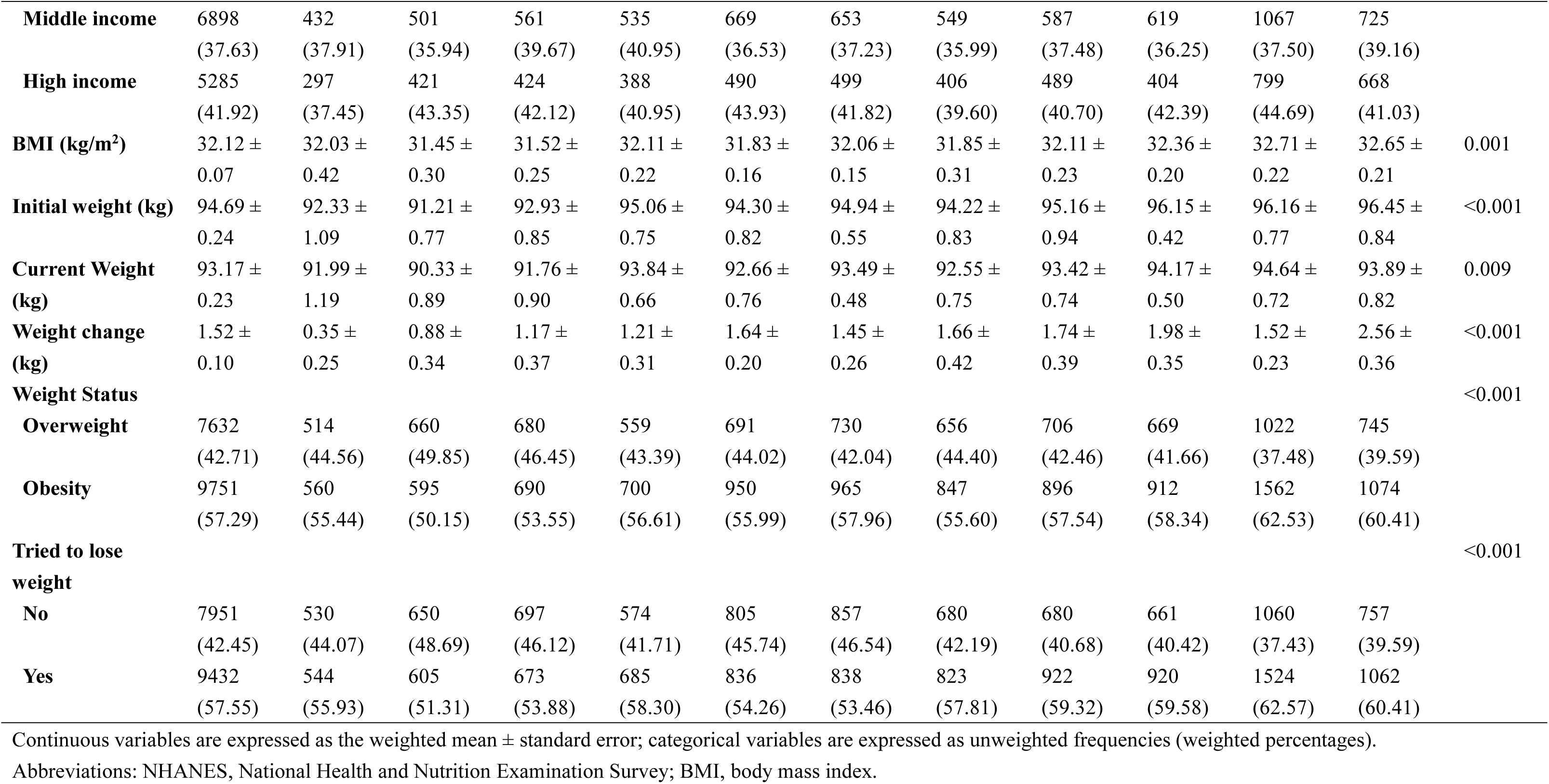
Characteristics of Participants by Survey Year.

### 3.2 Weight-loss attempts

Overall, 57.6% of adults with hypertension and overweight or obesity reported attempting to lose weight during the prior year (Table S2). Weight-loss attempts were more common among women than men (63.1% versus 52.5%; P<0.001). The prevalence of attempts also increased with educational attainment, from 45.9% among those with less than a high school education to 62.6% among those with a college education or above, and with income, from 52.2% in the low-income group to 62.2% in the high-income group (both P<0.001). Attempts were also markedly more common among adults with obesity than among those with overweight (67.5% versus 44.2%; P<0.001). By race and ethnicity, the prevalence of attempts ranged from 56.8% to 61.1%, with modest but statistically significant differences across groups (P=0.016; Table S2).

Over time, the overall prevalence of weight-loss attempts increased significantly, from 55.9% in 1999-2000 to 60.4% in 2021-2023 (Figure 2A; adjusted P for trend=0.002; Table S3). As shown in Figure 2B, this increase was more evident among men (P for trend=0.006) than women (P for trend=0.069). Significant increases were also observed among non-Hispanic White, non-Hispanic Black, and Mexican American participants (Figure 2C; P for trend=0.033, 0.020, and 0.011, respectively). By socioeconomic status, weight-loss attempts increased significantly in the low-income group, from 49.5% to 63.1% (Figure 2E; P for trend<0.001), but not in the middle- or high-income groups. When stratified by BMI category, attempts increased significantly among adults with obesity (Figure 2F; P for trend=0.007), whereas the increase among those with overweight did not reach statistical significance (P for trend=0.063). Additional trend estimates are provided in Table S3.

**Figure 2.**
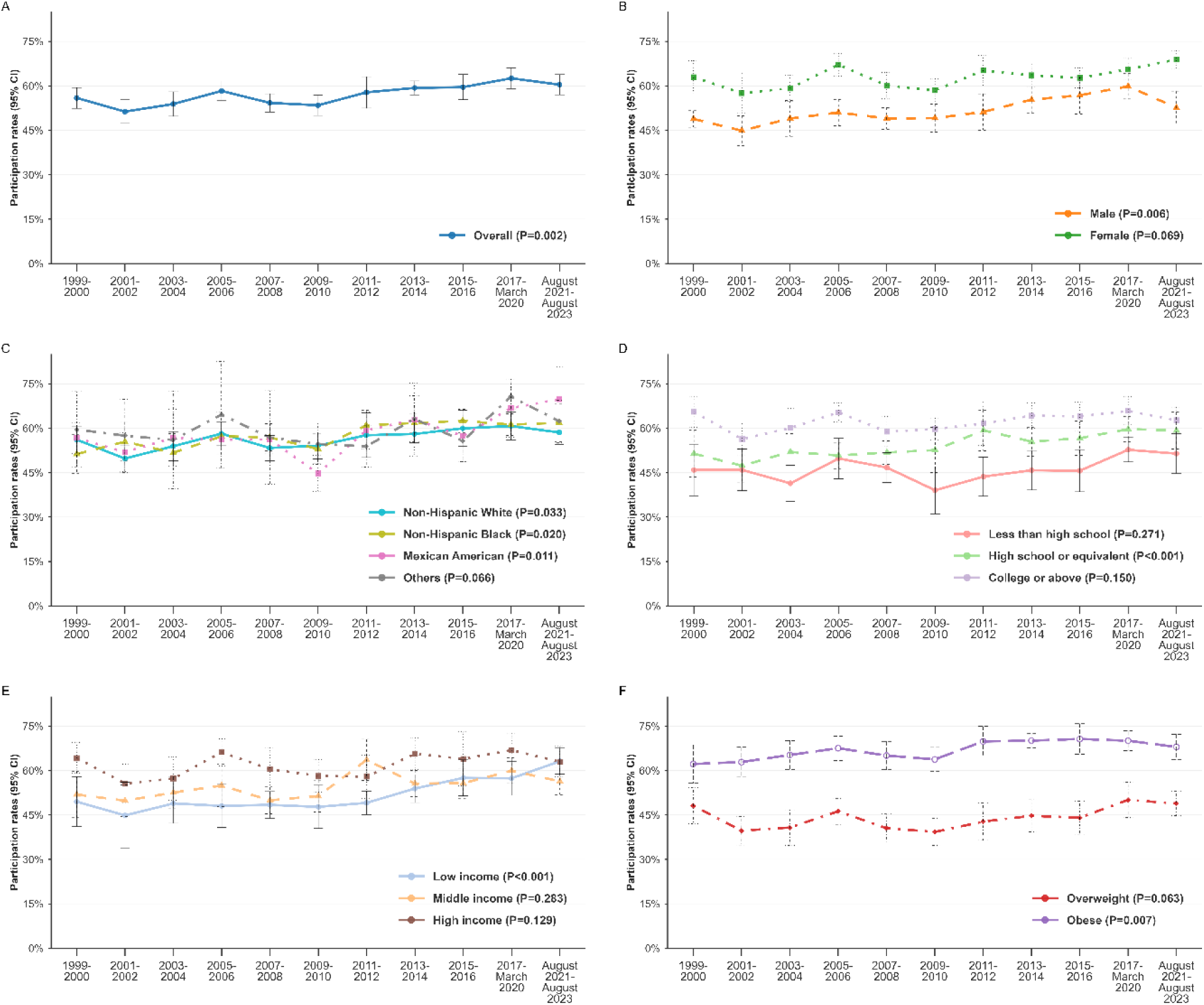
Trends in Weight Loss Attempts Among U.S. Adults with Hypertension: NHANES 1999–2023. (A) Overall; (B) Stratified by sex; (C) Stratified by race; (D) Stratified by Education Level; (E) Stratified by Family Income; (F) Stratified by BMI Categories. Data are weighted percentage. Error bars indicate 95% CIs.

### 3.3 Clinically meaningful weight loss

Among participants who reported attempting to lose weight, 33.4% achieved ≥5% weight loss and 14.7% achieved ≥10% weight loss over the prior year (Table S2). The prevalence of achieving ≥5% weight loss was similar in men and women (32.7% versus 34.0%; P=0.320) and did not differ significantly across racial and ethnic groups (range, 32.9%-34.8%; P=0.503). In contrast, significant differences were observed by income and BMI category. Participants in the low-income group were more likely than those in the high-income group to achieve ≥5% weight loss (38.1% versus 29.6%; P<0.001), and adults with obesity were substantially more likely than those with overweight to do so (40.0% versus 19.9%; P<0.001).

Over time, the prevalence of achieving ≥5% weight loss among those attempting to lose weight did not change significantly, increasing from 28.6% in 1999-2000 to 35.2% in 2021-2023 (Figure 3A; adjusted P for trend=0.976; Table S4). As shown in Figure 3B-E, this lack of temporal improvement was consistent across strata defined by sex, race and ethnicity, education, and income.

**Figure 3.**
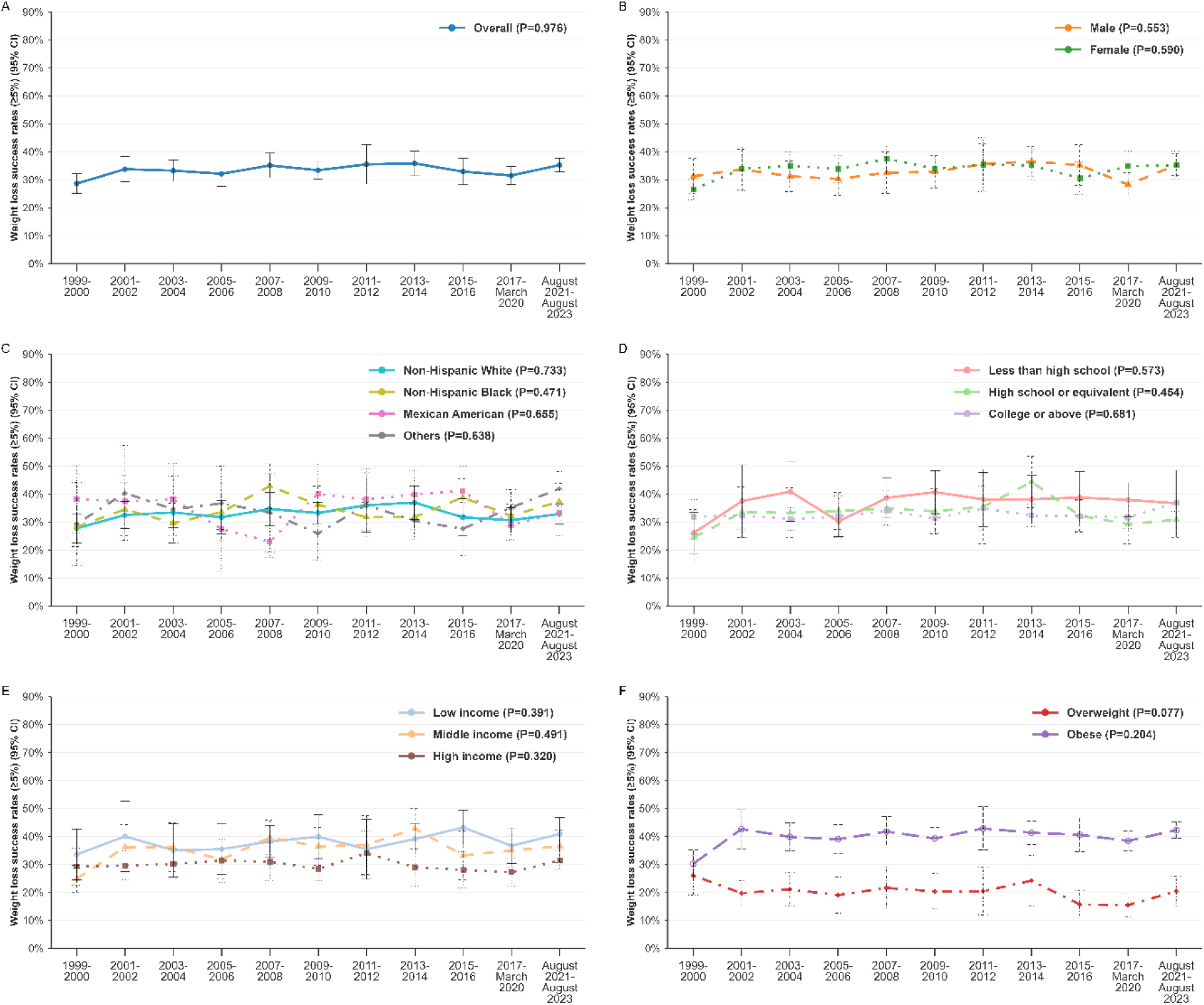
Trends in Achieving ≥5% Weight Loss Among U.S. Adults with Hypertension Who Attempted Weight Loss: NHANES 1999–2023. (A) Overall; (B) Stratified by sex; (C) Stratified by race; (D) Stratified by Education Level; (E) Stratified by Family Income; (F) Stratified by BMI Categories. Data are weighted percentage. Error bars indicate 95% CIs.

For the ≥10% threshold, achievement was more common among women than men (15.9% versus 13.2%; P=0.006) and differed by educational attainment (Table S2), with the highest prevalence among participants with less than a high school education (18.6%) compared with those with a high school education (14.1%) or college education or above (14.0%; P=0.004). Similar to the findings for ≥5% weight loss, achieving ≥10% weight loss was more common in the low-income group than in the high-income group (19.6% versus 11.9%; P<0.001) and in adults with obesity than in those with overweight (19.1% versus 5.6%; P<0.001; Table S2).

The prevalence of achieving ≥10% weight loss did not change significantly over time, increasing from 11.1% in 1999-2000 to 16.7% in 2021-2023 (Figure 4A; adjusted P for trend=0.174; Table S5). However, subgroup trends differed by BMI category (Figure 4F and Table S5). Among adults with overweight, achievement of ≥10% weight loss remained low and declined over time, from 8.6% to 5.1% (P=0.024). In contrast, among adults with obesity, the prevalence increased from 12.6% to 22.1% (P=0.024).

**Figure 4.**
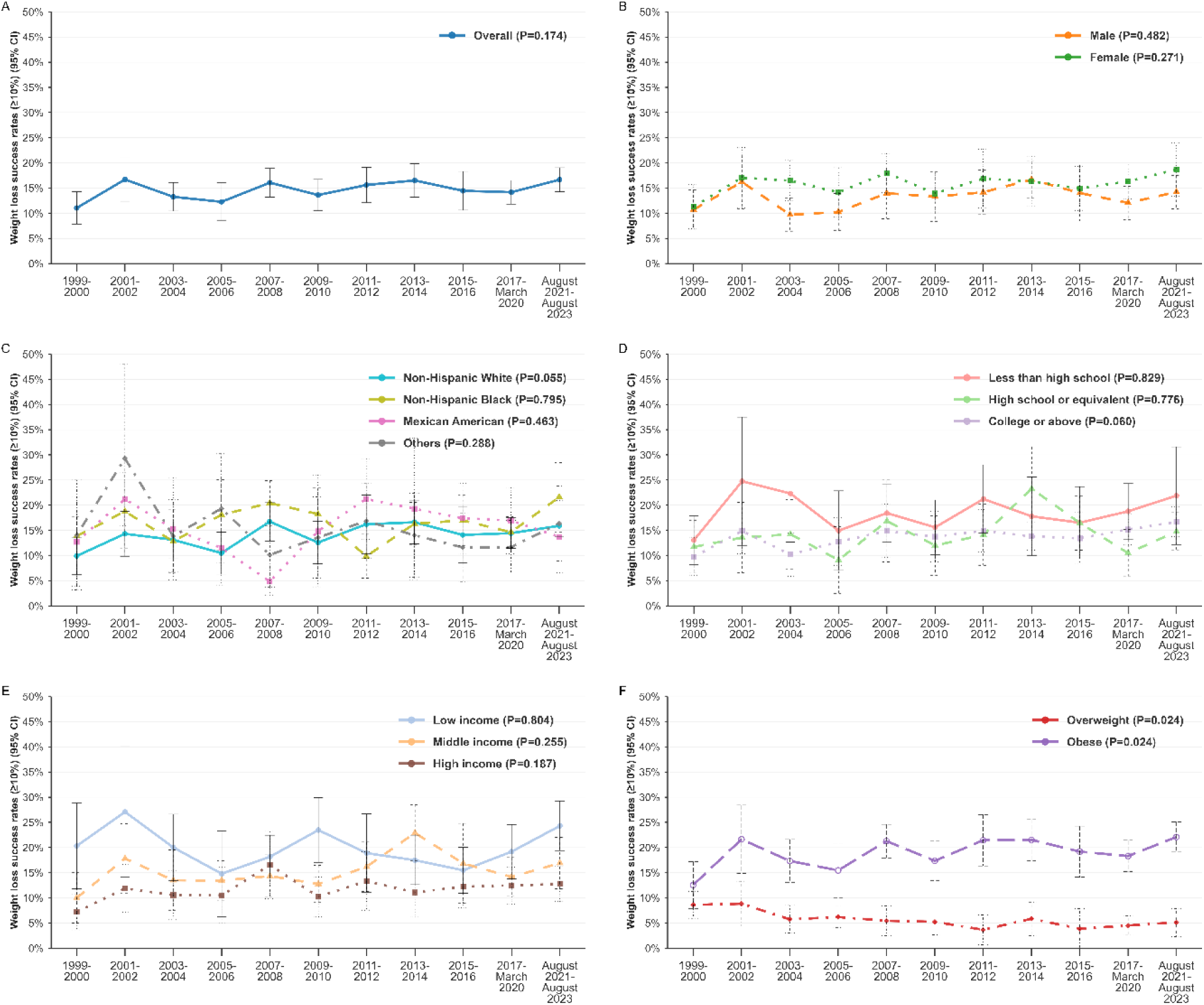
Trends in Achieving ≥10% Weight Loss Among U.S. Adults with Hypertension Who Attempted Weight Loss: NHANES 1999–2023. (A) Overall; (B) Stratified by sex; (C) Stratified by race; (D) Stratified by Education Level; (E) Stratified by Family Income; (F) Stratified by BMI Categories. Data are weighted percentage. Error bars indicate 95% CIs.

### 3.4 Weight-loss strategies

Among adults with hypertension who reported attempting to lose weight, the most commonly reported strategies during 1999-2000 through 2017-2020 were eating less food (65.3%), exercising (52.4%), drinking a lot of water (38.3%), eating less fat (36.8%), and choosing low-calorie foods (36.4%) (Figure 5A and Table S6). Other commonly reported dietary changes included eating more fruits, vegetables, or salads (28.2%), eating fewer carbohydrates (25.0%), eating less junk food or fast food (23.9%), eating less sugar, candy, or sweets (22.4%), and changing eating habits (21.7%; Figure 5A). Less frequently reported approaches included skipping meals (17.8%), using diet products (11.0%), following a special diet (8.1%), using a liquid diet formula (6.1%), joining a weight-loss program (6.1%), and taking nonprescription supplements (5.5%). Prescription diet pills (2.6%), laxatives or vomiting (1.0%), smoking (0.7%), and weight-loss surgery (0.4%) were rarely reported (Figure 5A).

**Figure 5.**
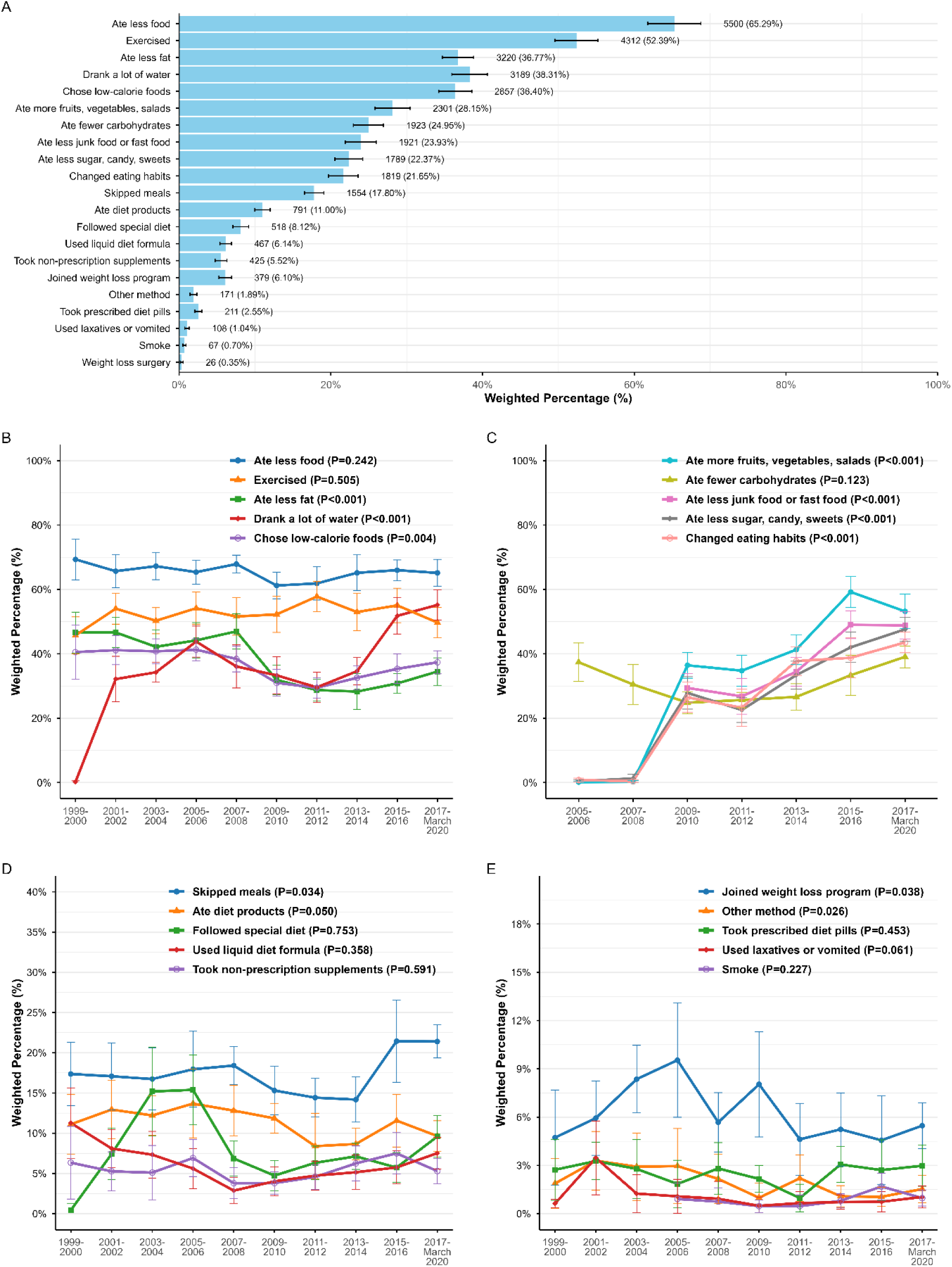
Trends and Prevalence of Weight Loss Measures Among U.S. Adults with Hypertension Who Attempted Weight Loss: NHANES 1999–2020. (A) Prevalence of weight loss measures; (B) Trends in Ate less food, Exercised, Ate less fat, Drank a lot of water, and Chose low-calorie foods; (C) Trends in Ate more fruits/vegetables/salads; Ate fewer carbohydrates; Ate less junk food or fast food; Ate less sugar/candy/sweets; and Changed eating habits; (D) Trends in Skipped meals; Ate diet products; Followed special diet; Used liquid diet formula; and Took non-prescription supplements; (E) Trends in Joined weight loss program; Other method; Took prescribed diet pills; Used laxatives or vomited; and Smoke. In panel A, data are presented as number of participants (weighted percentages). In panels B–E, data are presented as weighted percentages. Error bars indicate 95% confidence intervals.

Temporal trends in strategy use are shown in Figure 5B-E and Table S7. The prevalence of drinking a lot of water, eating more fruits, vegetables, or salads, eating less junk food or fast food, eating less sugar, candy, or sweets, and changing eating habits all increased significantly over time (all P<0.001; Figure 5B-C). Skipping meals also increased over time (P=0.034; Figure 5D). In contrast, eating less fat and choosing low-calorie foods declined significantly over the study period (P<0.001 and P=0.004, respectively; Figure 5B). The prevalence of eating less food and exercising remained stable over time (P=0.242 and P=0.505; Figure 5B). Among less commonly reported strategies, participation in a weight-loss program and reporting an “other” method increased over time (P=0.038 and P=0.026, respectively; Figure 5E), whereas prescription diet pill use showed no clear temporal trend (P=0.453).

### 3.5 Weight-loss strategies and successful weight loss

Associations between specific weight-loss strategies and successful weight loss are shown in Table S8. For achievement of ≥5% weight loss, eating less fat (OR, 1.19 [95% CI, 1.05-1.36]) and changing eating habits (OR, 1.26 [95% CI, 1.05-1.52]) were associated with higher odds of success in the multivariable model with backward stepwise selection. Reporting an “other” method (OR, 3.48 [95% CI, 2.17-5.60]), smoking (OR, 4.03 [95% CI, 2.05-7.93]), and weight-loss surgery (OR, 5.40 [95% CI, 1.57-18.53]) were also associated with higher odds of achieving ≥5% weight loss. In contrast, skipping meals was associated with lower odds of success (OR, 0.74 [95% CI, 0.63-0.86]; Table S8).

For achievement of ≥10% weight loss, changing eating habits remained positively associated with success (OR, 1.50 [95% CI, 1.22-1.86]). Reporting an “other” method (OR, 3.15 [95% CI, 1.99-4.99]), smoking (OR, 5.12 [95% CI, 2.37-11.07]), and weight-loss surgery (OR, 6.37 [95% CI, 2.16-18.79]) were likewise associated with higher odds of success. In contrast, use of diet products was inversely associated with achieving ≥10% weight loss (OR, 0.66 [95% CI, 0.48-0.90]; Table S8).

## 4. Discussion

In this nationally representative analysis spanning 1999-2023, we found that the proportion of US adults with hypertension and overweight or obesity who reported trying to lose weight increased substantially. This increase suggests growing awareness of weight management and broader uptake of lifestyle advice in a population at elevated cardiovascular risk. However, the central finding is that this greater engagement did not translate into better population-level outcomes. The proportion achieving clinically meaningful weight loss remained low and largely unchanged over time. Together, these findings point to a persistent gap between efforts to lose weight and successful weight reduction among adults with hypertension.

The increase in weight-loss attempts parallels prior national surveillance in the general US population [14], but our results show that greater engagement has not been accompanied by meaningful improvement in clinically significant weight loss among adults with hypertension. Attempts were more common among women than men and among adults with obesity than those with overweight, which may reflect differences in perceived risk, weight-related concerns, and the intensity of counseling delivered in clinical practice. Attempts were also more common among individuals with higher educational attainment and income, groups that may have greater health literacy, numeracy, and access to healthier food environments and opportunities for physical activity [22].

We also observed that the increase in weight-loss attempts over time was especially pronounced among low-income adults. This pattern may reflect expanded opportunities to initiate weight-loss efforts through policy and safety-net channels, including community-based nutrition and physical activity programs designed to reach low-income households [23–25]. However, greater engagement did not translate into higher rates of successful weight loss. This suggests that starting a weight-loss attempt may be more feasible than sustaining the behavioral changes needed to achieve clinically meaningful loss, particularly among low-income adults who face structural barriers such as food insecurity, limited access to affordable healthy foods, fewer safe environments for physical activity, and competing stressors that undermine long-term adherence [12, 18, 26, 27]. Without addressing these structural determinants, increasing attempts alone is unlikely to narrow the gap in outcomes.

A central finding of this study is that the prevalence of clinically meaningful weight loss did not improve over time despite a significant increase in weight-loss attempts. This pattern is consistent with the clinical reality that sustained weight reduction is difficult to achieve without structured and longitudinal support [28]. Intensive behavioral programs that provide concrete tools, frequent feedback, and ongoing follow-up generally produce the most reliable outcomes [29]. At the same time, newer anti-obesity pharmacotherapies have become increasingly important because they can substantially improve weight-loss outcomes when used appropriately as part of chronic obesity care [13]. In routine practice, however, many patients still rely on self-directed efforts while facing obesogenic environments, physiologic adaptations that favor weight regain, and uneven access to intensive behavioral treatment and pharmacotherapy [30, 31]. As a result, many patients may be unable to initiate or sustain effective treatment, contributing to the persistent gap between guideline recommendations and real-world outcomes [32].

The reported strategies for weight loss provide additional insight into this persistent intention-outcome gap. Eating less and exercising remained the most frequently reported strategies throughout the study period. Although these approaches are conceptually simple, “eat less and exercise more” is difficult to implement in practice; it requires patients to navigate obesogenic environments, overcome physiologic resistance to weight loss, and sustain long-term behavior change with limited feedback or reinforcement [30]. In our data, those achieving clinically meaningful weight loss were more likely to report specific and qualitative dietary changes rather than general caloric restriction. These findings suggest that the specificity and structure of dietary modification may distinguish successful from unsuccessful attempts, reinforcing the clinical view that generalized advice is less effective than strategies that translate into concrete, sustainable behaviors [33–35].

We also observed divergent trends in weight-loss success by BMI category. Among adults with obesity, the prevalence of achieving 10% weight loss increased substantially over time, whereas it remained low and declined among adults with overweight. Clinically, providers may be more likely to pursue intensive counseling and treatment escalation once patients reach the obesity range, when the perceived urgency is greater and conversations shift from prevention to active treatment [36, 37]. Patients with obesity may also be more likely to receive referrals to structured programs, consideration for pharmacotherapy, or evaluation for metabolic surgery [38]. By contrast, individuals with overweight may receive briefer, less structured counseling, and weight management may be framed as general lifestyle advice rather than as treatment for a chronic condition requiring a plan and follow-up.

We also observed consistently low use of evidence-based, high-intensity interventions. Only 6.1% of individuals attempting weight loss reported participation in a weight-loss program. Prescription anti-obesity medication use was 2.6%, and metabolic surgery was reported by 0.4%. The gap between interventions with demonstrated efficacy and those actually used in practice remains substantial. In the absence of organized programs, pharmacotherapy, or surgical referral pathways, many patients appear to rely on broadly accessible but variably effective approaches such as eating less, increasing exercise, drinking more water, or skipping meals. These efforts are often undertaken without a structured plan, objective monitoring, or timely escalation of treatment. Some commonly used behaviors may even be counterproductive, such as meal skipping [39, 40]. Improvements in dietary quality may be beneficial, but for many patients they may still be insufficient to achieve and sustain clinically meaningful weight loss without additional longitudinal support [41].

The temporal trends in strategy adoption offer both encouraging and cautionary signals. The increased prevalence of dietary quality measures over time, including greater consumption of fruits and vegetables and reduced intake of sugar and fast food, suggests that public health messages emphasizing healthy eating patterns have been absorbed [8, 42]. In contrast, the proportion reporting exercise as a weight-loss strategy remained relatively stable. Whether this stability reflects persistent structural barriers, inadequate clinical counseling, or other constraints is unclear [43], but it points to an enduring opportunity for both public health and clinical intervention. At the same time, eating less fat and switching to foods with lower calories declined over time. Reduced use of these strategies may partly help explain why clinically meaningful weight-loss success did not improve significantly despite increasing weight-loss attempts.

Several findings require careful interpretation. The strong association between smoking and weight-loss success should not be misconstrued as support for tobacco use as a weight-loss strategy. Smoking can reduce body weight through appetite suppression and increased metabolic rate, but its cardiovascular harms, including increased blood pressure, are well established [44–46]. In adults with hypertension, smoking represents net harm regardless of any association with weight loss. The observed positive relationship should therefore be interpreted as an epidemiologic association rather than a target for intervention. Similarly, the strong association between metabolic surgery and weight-loss success is consistent with the established efficacy of surgical approaches, but the very low prevalence of surgery indicates major barriers to access. For individuals with hypertension and obesity who do not achieve adequate weight loss with lifestyle modification and pharmacotherapy, surgery is an evidence-based option that can produce durable weight loss and improve blood pressure and cardiometabolic risk [47]. The low use of surgery in this population may therefore be interpreted as a system-level access gap rather than an absence of clinical need.

Looking forward, the emergence of highly effective anti-obesity pharmacotherapies has the potential to change population trajectories. Agents such as GLP-1 receptor agonists and related incretin-based therapies can produce substantial weight loss, approaching that achieved with metabolic surgery in some patients [48]. However, their impact will depend on factors that have historically limited the reach of effective obesity treatment, including access, affordability, equitable prescribing, and sustained use [49]. Discontinuation due to cost, adverse effects, or inadequate longitudinal support may limit long-term effectiveness, and weight regain commonly occurs after treatment cessation [50]. The promise of pharmacotherapy is most likely to be realized when these agents are delivered within comprehensive care models that include behavioral support, structured follow-up, and equitable access.

These results carry direct implications for hypertension care. Many adults with hypertension and excess weight are already trying to lose weight, yet sustained success remains uncommon at the population level. Advice from healthcare professionals alone is unlikely to close the gap between intention and outcome [51]. Instead, weight management may need to be operationalized as a chronic-care process alongside blood pressure management, with explicit goals, routine monitoring, longitudinal support, and treatment escalation when response is inadequate [52]. Health systems may consider integrating dietitians, health coaches, and behavioral programs into routine hypertension care; establishing clear referral pathways to evidence-based interventions; and using registries or follow-up systems to track progress and intensify treatment when needed [53, 54]. Current obesity guidance increasingly supports multidisciplinary, team-based treatment models [55]. Health systems may benefit from integrating dietitians, pharmacists, advanced practice clinicians, health coaches, and physicians with expertise in obesity medicine into routine hypertension care, while establishing clear referral pathways to evidence-based behavioral treatment, pharmacotherapy, and metabolic surgery. At the policy level, improving affordability and coverage for intensive behavioral programs, pharmacotherapy, and surgery may be essential to enable effective care at scale [49].

Future research should clarify why weight-loss success has remained unchanged despite increasing attempts. Priorities include evaluating the use, persistence, and equity of newer anti-obesity pharmacotherapies and structured behavioral programs in this population, as well as determining whether weight loss achieved in routine practice is durable and associated with improvements in blood pressure control and medication burden. Additional work should test care delivery models that integrate obesity treatment into hypertension clinics, including team-based approaches, telehealth-enabled behavioral treatment, and stepwise treatment escalation. Implementation studies that evaluate scalable strategies in real-world settings, particularly those that address social determinants and resource constraints, will be important for translating guideline recommendations into population-level improvement.

Several limitations of this study should be acknowledged. Weight-loss attempts, strategies, and weight change were based on self-report and may be affected by recall bias or social desirability bias. Although the consistency of our findings with objectively measured BMI trends provides some reassurance, self-reported weight may underestimate true weight, potentially inflating calculated weight loss and biasing success estimates. The observational, cross-sectional design precludes causal inference regarding associations between strategies and success. Unmeasured confounding, including prior weight-loss attempts, motivation level, and concurrent medical conditions, may influence these relationships. The temporal availability of certain weight-loss strategy items varied across cycles. The small numbers of participants reporting certain strategies, such as weight-loss surgery and smoking for weight loss, yielded imprecise estimates reflected in wide confidence intervals. Our definition of weight-loss success captures clinically meaningful thresholds but does not account for durability of loss beyond one year or the distribution of weight loss. Finally, NHANES does not include institutionalized individuals, and findings may not generalize to all US adults with hypertension.

## Conclusions

In this nationally representative analysis of US adults with hypertension and excess weight from 1999 through 2023, the proportion reporting weight-loss attempts increased significantly, yet the proportion achieving clinically meaningful weight loss did not improve. These findings suggest that motivation and engagement have increased, but current approaches do not reliably translate effort into success at the population level. Closing this intention-to-outcome gap will require treating obesity as a chronic condition within hypertension care, with structured longitudinal support, equitable access to intensive behavioral programs, appropriate use of pharmacotherapy and metabolic surgery, and consistent follow-up with treatment escalation when needed.

## List of abbreviations

AHA: American Heart Association
ACC: American College of Cardiology
BMI: body mass index
BP: blood pressure
CI: confidence interval
ESC: European Society of Cardiology
ISH: International Society of Hypertension
NHANES: National Health and Nutrition Examination Survey
OR: odds ratio

## Declarations

Not applicable

## Consent for publication

Not applicable

## Availability of data and materials

The datasets generated and analysed during the current study are available in the NHANES, https://wwwn.cdc.gov/nchs/nhanes/nhefs/default.aspx

## Authors’ contributions

G.X. and R.T. conducted NHANES data collection, collation, and preliminary cleaning. C.H. provided clinical insights, refined the research framework, and advised on outcome interpretation. M.S., Y.C., and J.L. contributed clinical expertise in weight-loss strategy evaluation and indicator analysis and revised the clinical content. T.H. and L.L. performed statistical analysis of the NHANES dataset, ensuring result reliability. Y.C. reviewed the manuscript and data and provided critical suggestions for revision. All authors gave their consent for authorship and inclusion in this manuscript. No AI or language models were used beyond basic tools.

## Acknowledgements

We acknowledge NHANES for dataset support.

